# Automated 360-degree goniophotography with the NIDEK Gonioscope GS-1 for glaucoma

**DOI:** 10.1101/2022.06.22.22276741

**Authors:** Chisom T. Madu, Taylor Phelps, Joel S. Schuman, Ronald Zambrano, Ting-Fang Lee, Joseph Panarelli, Lama Al-Aswad, Gadi Wollstein

**Affiliations:** Department of Ophthalmology, NYU Langone Health, NYU Grossman School of Medicine, New York, NY, United States of America; Department of Neuroscience and Physiology, NYU Langone Health, NYU Grossman School of Medicine, New York, NY, United States of America; Departments of Biomedical Engineering and Electrical & Computer Engineering, New York University Tandon School of Engineering, Brooklyn, NY, United States of America; Center for Neural Science, NYU College of Arts and Sciences, New York, NY, United States of America

**Keywords:** Anterior chamber angle, gonioscopy, automated imaging, diagnostic testing, iridocorneal angle

## Abstract

**Purpose:** To evaluate the reliability of the NIDEK Gonioscope GS-1 when used to grade the iridocorneal angle in a clinical setting.

**Methods:** A total of 20 participants (37 eyes) who were 18 or older and had glaucoma or were glaucoma suspects were enrolled from the NYU Langone Eye Center and Bellevue Hospital. During their usual ophthalmology visit, they were consented for the study and underwent 360-degree goniophotography using the NIDEK Gonioscope GS-1. Afterwards, the three ophthalmologists separately examined the images obtained and determined the status of the iridocorneal angle in four quadrants using the Shaffer grading system. Physicians were masked to patient names and diagnoses. Inter-observer reproducibility was determined using Fleiss’ kappa statistics.

**Results:** The interobserver reliability using Fleiss’ statistics was shown to be significant between three glaucoma specialists with fair overall agreement (Fleiss’ kappa: 0.266, p<.0001) in the interpretation of 360-degree goniophotos. Automated 360-degree goniophotography using the NIDEK Gonioscope GS-1 have quality such that they are interpreted similarly by independent expert observers. This indicates that angle investigation may be performed using this automated device and that interpretation by expert observers is likely to be similar.

**Conclusion:** Images produced from automated 360-degree goniophotography using the NIDEK Gonioscope GS-1 are similarly interpreted amongst glaucoma specialists, thus supporting use of this technique to document and assess the anterior chamber angle in patients with, or suspected of, glaucoma and iridocorneal angle abnormalities.

**Précis:** Automated 360-degree goniophotography using the NIDEK Gonioscope GS-1 is a reasonable tool for anterior chamber angle assessment.

## Background

Gonioscopy is a vital part of the ophthalmologic examination, as it allows clinicians to evaluate the iridocorneal angle of the eye. The iridocorneal angle contains the orifice to the conventional drainage pathway of the aqueous humor from the anterior chamber into the Schlemm canal. Narrow, occludable angles can put patients at risk for acute angle closure glaucoma due to obstruction of the route of the fluid into the trabecular meshwork. Accumulation of debris or inflammatory cells can also lead to obstructions of the drainage system and consequent increase in intraocular pressure. Gonioscopy allows clinicians to assess the access to and width of the angle as well as look for structural deformation, pigmentation, inflammation, or accumulation of extraneous materials.

Although gonioscopy plays an important role in glaucoma diagnosis and treatment, repeatability and reliability are user-dependent when a conventional goniolens is used. During examination, the goniolens must be manipulated to a significant degree to obtain full visualization of the iridocorneal angle. While it is considered the gold standard for assessment of the iridocorneal angle, gonioscopy can be difficult to perform[1]. The evaluation of the width of the iridocorneal angle is subjective and requires clinical experience[1]. As such, there is inter-operator variability in the results obtained using this method.

Automated 360-degree goniophotography (Gonioscope GS-1, NIDEK, Gamagori, Japan) captures iridocorneal angle images using a 16-face multi-mirror optical gonioprism and a built-in image sensor. Through the use of an intrinsic rotator unit, a colored circumferential image of the iridocorneal angle and its peripheral area is captured in a single examination. If demonstrated that the images provided from the NIDEK GS-1 are similarly interpreted by masked glaucoma specialists, then it could be used as an effective screening tool to determine which patients may need a formal evaluation for angle abnormalities, including narrow angles, structural abnormalities, pigmentation, inflammation, debris and masses, thus among other things, preventing angle closure events and blindness.

The interobserver agreement between physicians for the grading of gonio-images produced by the NIDEK GS-1 has previously been reported by Teixeira et al. and Matsuo et al. using Shaffer and Scheie grading systems respectively[2, 3]. Teixeira et al. found that there was moderate agreement (Fleiss’ kappa: 0.48) between a glaucoma specialist and an ophthalmology resident in the assessment of 47 patients (88 eyes) using the NIDEK GS-1[2]; while Matsuo et al. observed slight agreement (Fleiss’ kappa: 0.17) between three glaucoma specialists and two general ophthalmologists, and fair agreement (Fleiss’ kappa: 0.31) when comparing between only the three glaucoma specialists, in the assessment of 35 patients (35 eyes) using the NIDEK GS-1[3]. The purpose of this study is to similarly determine the agreement between three glaucoma specialists in the grading of the iridocorneal angle using the Shaffer scoring system in automated 360-degree goniophotography images among a cohort of patients.

## Methods

### Overview of Study Design

This study was conducted in compliance to the IRB, GCP guidelines, and applicable NYU Langone Health and federal regulatory requirements.

Goniophotos of glaucoma and glaucoma suspect subjects were taken by the NIDEK GS-1 and independently assessed by three glaucoma specialists (LAA, JSS, and JP) using the Shaffer grading system. Gradings for the width of the irideocorneal angle were then compared between physicians to determine reliability. The Department of Ophthalmology, NYU Langone Health, was responsible for test data preparation, data acquisition, and analyses.

### Study subjects

Study subjects were screened in advance and then approached during their routine ophthalmology visits for written consent. A total of 22 subjects were enrolled in the study. Of those enrolled, two subjects failed to return to receive the experimental intervention and were subsequently lost to follow-up. 20 subjects and 37 eyes were included in the study. All subjects were 18 years or older and diagnosed with glaucoma or determined to be a glaucoma suspect. A diagnosis of glaucoma was based on the identification of clinical glaucomatous retinal nerve fiber layer (RNFL) defects and optic nerve head (ONH) abnormalities, including global rim thinning, rim notching, disc hemorrhage, or two consecutive reliable visual field tests with glaucoma hemifield test (GHT) showing a mean deviation outside normal limits. A diagnosis of glaucoma suspect was based on having glaucomatous optic neuropathy, as described for the diagnosis of glaucoma, and/or having ocular hypertension (IOP > 21 mm Hg) with a most recent visual field test with GHT showing a mean deviation within normal limits. Criteria for exclusion included the presence of corneal opacities, such as scars and edema, pregnancy or planning to be pregnant, and an inability to fixate gaze.

### Gonioscope GS-1 imaging

During their usual ophthalmology visit, subjects underwent conventional gonioscopy, ultrasound biomicroscopy, and gonioscopy with the NIDEK Gonioscope GS-1. The NIDEK Gonioscope GS-1 was only used on eligible patients who were consented for the study. A coupled gel was applied to the prism tip prior to using the device. This gel reduces the refraction angle between the patient’s cornea and the prism and moderates the variation in each patient’s corneal shape. Because the multi-mirror prism touches the surface of the eye it carries a risk of infection. We performed high level disinfection or low temperature sterilization (EOG sterilization) after any use of the prism to reduce this risk. Additionally, all exposed surfaces near the eye, as well as the chin rest and forehead rest of the instrument, were cleaned with 70% isopropyl alcohol before participants are examined. The multi-mirror prism is made from cycloolefin polymer (COP) resin. In vitro cytotoxicity assays found that concentrations of up to 100% COP did not obstruct colony formation of V79 Chinese hamster lung cells. Ocular irritation tests showed that COP does not irritate the cornea, iris, or conjunctiva of rabbits.

Although the NIDEK Gonioscope GS-1 is not currently an FDA-approved device, it is a non-significant risk device under the criteria of 21 CFR 812.3 (m):

1. It is NOT intended as an implant and does NOT present a potential for serious risk to the health, safety, or welfare of a subject
2. It is NOT purported or represented to be for a use in supporting or sustaining human life and does NOT present a potential for serious risk to the health, safety, or welfare of a subject
3. It is for a use of substantial importance in diagnosing, curing, mitigating, or treating disease, or otherwise preventing impairment of human health and does NOT present a potential for serious risk to the health, safety, or welfare of a subject

### Evaluation of GS-1 goniophotos

NIDEK GS-1 goniophotos were independently examined by three glaucoma specialists from the NYU Langone Eye Center and board-certified by the American Board of Ophthalmology (ABOP). Response forms and goniophotos were sent to observers by e-mail, who then completed diagnostic evaluations of all images. The width of the iridocorneal angle was measured using the Shaffer grading system and ranked from 0-4[4]. A grade of 0 is defined as a “closed” angle with no visible angle structures, a grade of 1 as “extremely narrow” with visibility up to Schwalbe’s line, a grade of 2 as “narrow” with visibility up to the trabecular meshwork, a grade of 3 as “open” with visibility up to the scleral spur, and a grade of 4 as “wide open” with visibility up to the ciliary body band[4]. The three observers were masked to patient names and diagnoses.

### Statistical analysis

Repeatability between glaucoma specialists was analyzed for the assessment of Shaffer’s angle from GS-1 goniophotos[2, 3, 5]. Interobserver agreement was measured using Fleiss’ kappa statistics and verified through the calculation of 95% confidence intervals[6, 7]. Kappa statistics were categorized as either poor (<0.20), fair (0.20–0.40), moderate (0.40–0.60), substantial (0.60– 0.80), or excellent (>0.80), following the guideline established by Altman[8]. Statistical analysis was carried out using GraphPad Prism version 8 (GraphPad, La Jolla, CA) and SPSS Statistics version 25 (IBM Corporation). Demographic and clinical characteristics were expressed as the mean and standard deviation for continuous variables, or by number and percentage for discrete variables.

## Results

In total, 37 eyes from 20 patients were included in the study. The demographic and clinical characteristics of the study population are detailed in **Table 1**. Subjects had a mean age of 56.4 ± 12.5 years. 70% of the subjects were female (n = 14).

**Table 1.**
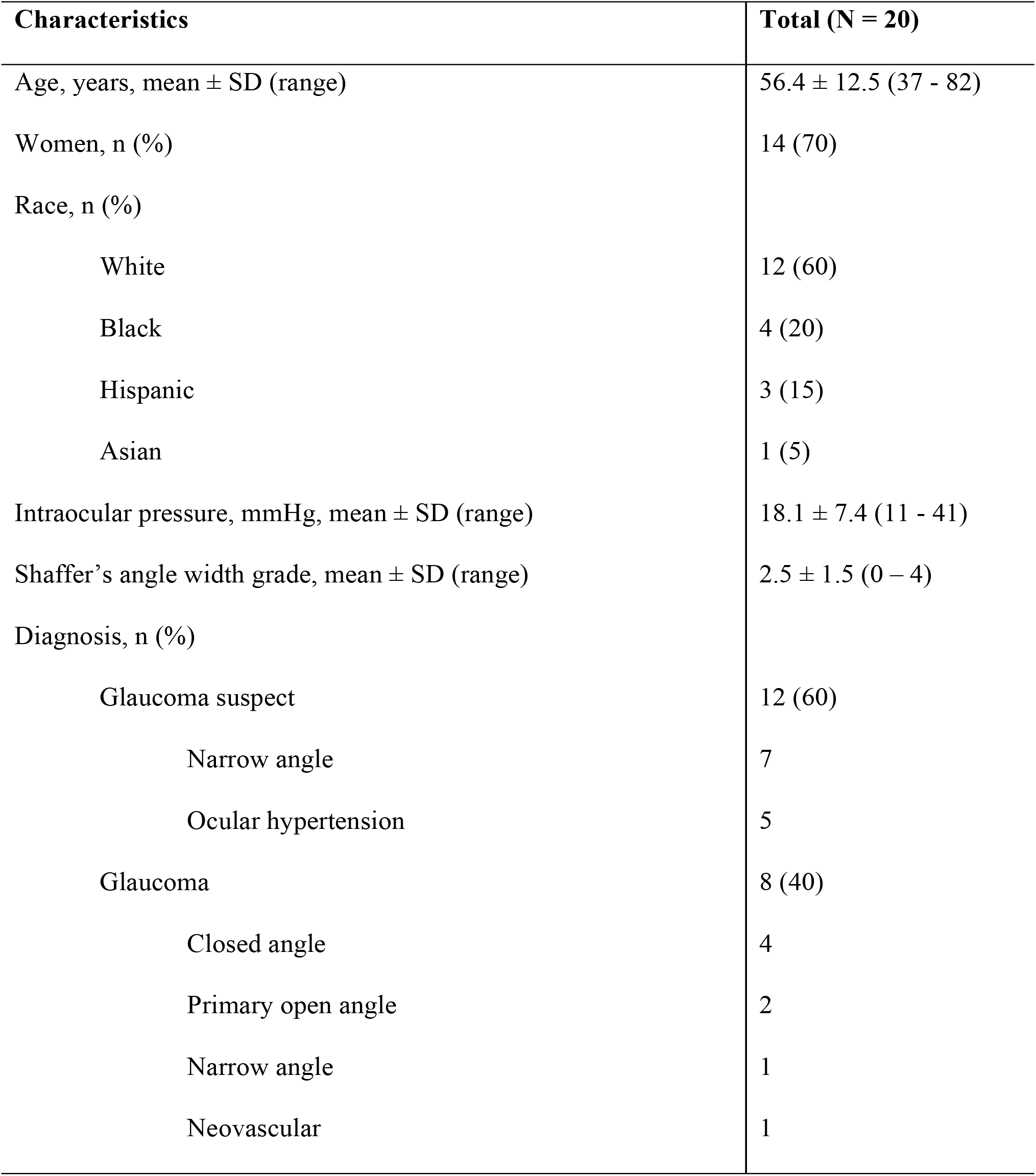
Demographic and Clinical Characteristics of the Study Population.

| Characteristics | Total (N = 20) |
| --- | --- |
| Age, years, mean ± SD (range) | $56.4 \pm 12.5$ (37 - 82) |
| Women, n (%) | 14 (70) |
| Race, n (%) |  |
| White | 12 (60) |
| Black | 4 (20) |
| Hispanic | 3 (15) |
| Asian | 1 (5) |
| Intraocular pressure, mmHg, mean ± SD (range) | $18.1 \pm 7.4$ (11 - 41) |
| Shaffer's angle width grade, mean ± SD (range) | $2.5 \pm 1.5$ (0 - 4) |
| Diagnosis, n (%) |  |
| Glaucoma suspect | 12 (60) |
| Narrow angle | 7 |
| Ocular hypertension | 5 |
| Glaucoma | 8 (40) |
| Closed angle | 4 |
| Primary open angle | 2 |
| Narrow angle | 1 |
| Neovascular | 1 |

### Interobserver agreement in angle assessment

**Table 2** shows the values of interobserver agreement in Shaffer angle width grading of NIDEK GS-1 goniophotos among the three glaucoma specialists overall as well as for each of the four quadrants imaged. The Fleiss’ kappa coefficient of reliability among all observers and across all quadrants was 0.266 (fair agreement). The individual Fleiss’ kappa coefficients among observers for superior, nasal, inferior, and temporal regions were found to be 0.257, 0.294, 0.261, and 0.246 respectively (all fair agreement).

**Table 2.** Interobserver Agreement of Shaffer Angle Width Grading in the Analysis of NIDEK GS-1 Goniophotography.

|  | Overall |  | Superior | Nasal | Inferior | Temporal |
| --- | --- | --- | --- | --- | --- | --- |
|  | Fleiss' kappa coefficient | Landis-Koch score | Fleiss' kappa coefficient | Fleiss' kappa coefficient | Fleiss' kappa coefficient | Fleiss' kappa coefficient |
| **All three observers** | 0.266** | *Fair Agreement* | 0.257** | 0.294** | 0.261** | 0.246** |
| **Observers 1 & 2** | 0.369** | *Fair Agreement* | 0.356** | 0.481** | 0.363** | 0.288* |
| **Observers 2 & 3** | 0.155* | *Weak Agreement* | 0.125 | 0.203 | 0.142 | 0.143 |
| **Observers 1 & 3** | 0.254* | *Fair Agreement* | 0.268* | 0.217* | 0.258* | 0.26* |

\*P-value < 0.05
\*\*P-value < 0.001

## Discussion

Images of the iridocorneal angle obtained by the NIDEK Gonioscope GS-1 were independently graded for angle width and analyzed by physicians to determine interobserver reliability using Fleiss’ statistics. The kappa coefficient for the overall reliability of grading 360-degree goniophotos in this study was found to be 0.266, which is in agreement with results from similar studies by Teixeira et al. and Matsuo et al. who reported overall kappa coefficients of 0.31 and 0.17, respectively. Findings between the three observers in this study demonstrated fair agreement across superior, nasal, inferior, and temporal quadrants. We did not find any correlation between image quadrant and interobserver agreement.

The clinical standard for the assessment of the iridocorneal angle is conventional gonioscopy conducted using a handheld goniolens. The pooled interobserver agreement between five glaucoma specialists in measuring the width of the iridocorneal angle using conventional gonioscopy with a binary scale (narrow or open) was found to be substantial across two visits (Fleiss’ kappa: 0.66, 0.69)[9]. Although advantageous because of the ability of the physician to manipulate the goniolens for static and dynamic observation of the anterior segment, conventional gonioscopy has no capacity for visual documentation, and thus, is not optimized for long-term patient follow-up or remote assessment. Furthermore, the practice of conventional gonioscopy is subjective and heavily operator dependent. Factors that can impact the findings of the physician include corneal pressure, lighting conditions, angle pigmentation, and iris convexity[3]. Matsuo et al. suggest that a combined approach of using both 360-degree goniophotography and conventional gonioscopy techniques for general screening and then the thorough examination of suspicious cases respectively may be effective[3].

Due to the time and skill required to obtain an accurate gonioscopic image using a conventional slit lamp, there has been interest in developing alternative methods to obtain quick, accurate, and reliable images of the iridocorneal angle. These alternative methods have involved the use of both new and existing technologies. For instance, currently available 3D Corneal and Anterior Segment Optical Coherence Tomography (3D CAS-OCT, CASIA, Tomey Corp, Nagoya, Japan) has been used for this purpose. A study investigating the utility of 3D CAS-OCT in evaluating anterior segment parameters found a high degree of intra-grader repeatability, inter-grader repeatability, and correlation with conventional gonioscopy[10]. Assessment of the angle may also be performed using conventional spectral domain (SD) and swept source (SS) OCT. Swept source has the advantage of greater tissue penetration and therefore a view of the angle superior to SD-OCT.

With the goal of someday replacing conventional gonioscopy, some imaging devices have already been developed to specifically assess anterior segment parameters. The Pentacam-Scheimpflug camera (OCULUS, Inc, Arlington, WA) uses the Scheimpflug optical principle to photograph parts of the anterior segment that are not directly in line with the camera’s aperture[11]. The camera takes up to 50 slit images of the anterior segment in 2 seconds and uses software to construct a 3D image[11]. The benefit of this technology is that it requires little operator experience and begins scanning automatically once proper alignment has been achieved[11]. However, Pentacam-Scheimpflug only quantitatively estimates the width of the angle and does not directly visualize it. Orbscan Scanning-Slit Topography (Bausch + Lomb Inc., Bridgewater, NJ) is a technology that scans the entire surface of the cornea, the iris, and the lens using a slit-scanning beam[11]. By mapping both the posterior surface of the cornea and the iris, Orbscan calculates an estimation of the iridocorneal angle[11]. However, like Pentacam-Scheimpflug, it does not directly visualize the angle. Ultrasound biomicroscope can obtain images of the iridocorneal angle and has been used for decades in the form of A- and B-scans[11]. However, to obtain an image, it requires direct contact with the eye, which can be uncomfortable for the patient and is highly operator dependent[11]. The NIDEK GS-1 has the capability of being able to directly image the iridocorneal angle with minimal patient discomfort, as well as true color imaging of the trabecular meshwork and angle structures.

Limitations of this study include the small size of the sample population as well as having an uneven distribution of sample population characteristics. Most subjects included in the study were white (60%) and female (70%). A more diverse population would give the findings of this study greater clinical applicability. Additional research to determine the level of agreement between 360-degree goniophotography and conventional goniolens gonioscopy in iridocorneal angle assessment is needed to better understand the clinical utility of this imaging technique and its potential role in the future.

This study confirms the results of previous findings regarding the practicality and reproducibility of goniophotography. In conclusion, images produced from automated 360-degree goniophotography using the NIDEK Gonioscope GS-1 are similarly interpreted amongst glaucoma specialists. This suggests that 360-degree goniophotography may be a useful imaging technique in screening and assessing the anterior chamber angle in patients with, or suspected of, glaucoma and iridocorneal angle abnormalities.

## Data Availability

Data cannot be shared publicly because of patient protected health information. Data are available from the NYU Langone Health Institutional Data Access / Ethics Committee (contact via 212-263-4110) for researchers who meet the criteria for access to confidential data.

---

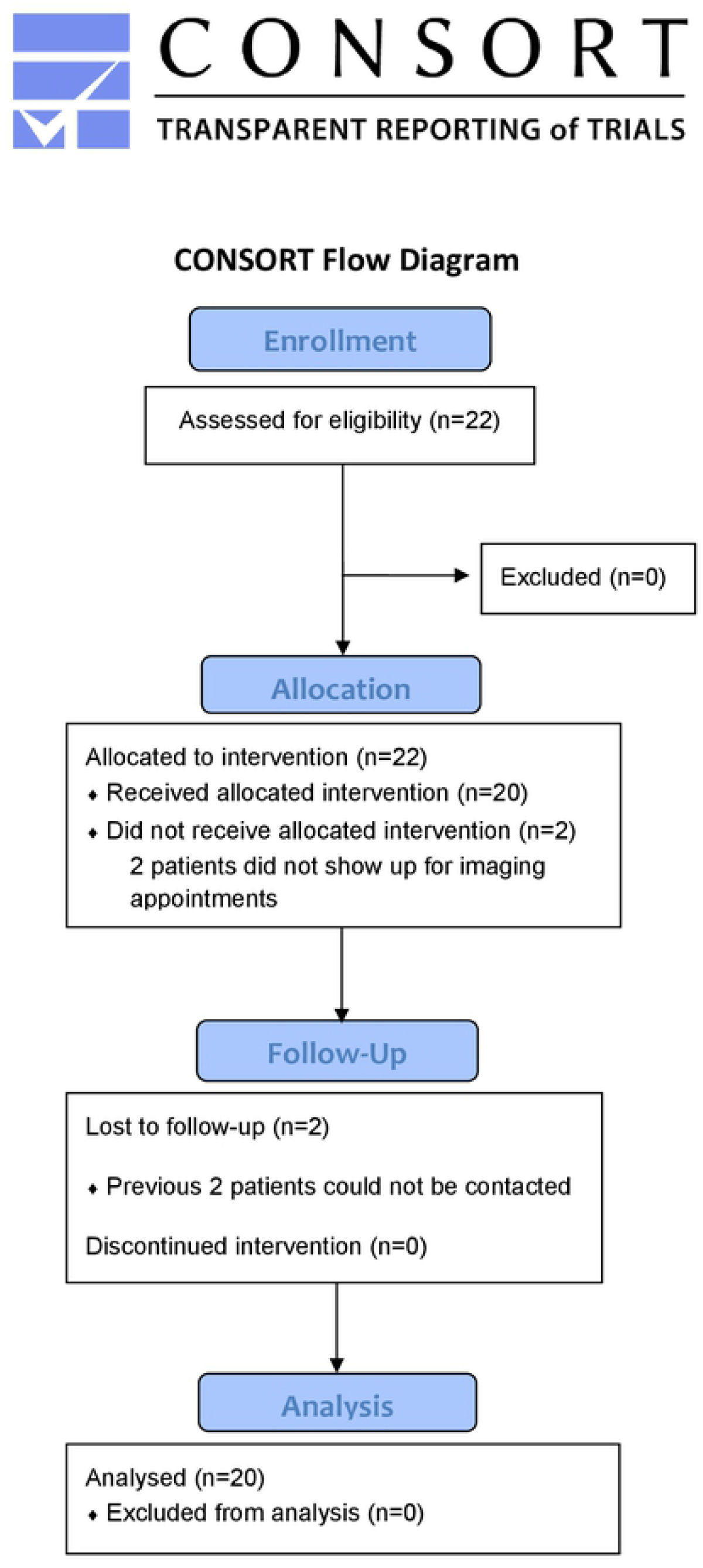
CONSORT Flow Diagram

